# The potential benefits of delaying seasonal flu vaccine selections: a retrospective modeling study

**DOI:** 10.1101/2023.04.26.23289161

**Authors:** Kyueun Lee, Katherine Williams, Janet Englund

**Affiliations:** Department of Pharmacy, School of Pharmacy, University of Washington, Seattle, Washington, USA; Department of Family Medicine, School of Medicine, University of Pittsburgh, Pittsburgh, Pennsylvania, USA; Department of Infectious Diseases, Seattle Children’s Hospital, University of Washington, Seattle, Washington, USA

**Keywords:** Seasonal flu, vaccine, mathematical model

## Abstract

**Backgrounds and Purpose:** Antigenic match between selected vaccine virus and circulating virus crucial to achieve high vaccine effectiveness for seasonal flu. Due to the time-consuming process of producing eggs, vaccine candidate viruses are currently selected 5-6 months ahead of the flu season. New non-egg-based vaccine production technologies have emerged with the potential to improve production efficiency and to revise current vaccine formulation schedules. In this study, we aim to 1) identify the past flu seasons where the opportunity to improve vaccine decision existed if rapid vaccine production were available and to 2) quantify the impact of revising the current vaccine decision schedule, where new vaccine production technologies allow more time for specimen collection prior to vaccine virus selection.

**Methods:** We extracted the trend in the viral activity of season-predominant strain in three data points: when vaccine decision was made, in between vaccine decision and season starts, and after season starts. Between 2012 and 2020, we first identified the past flu seasons where the season-dominant strains presented increasing activity only after vaccine decisions had already been made in February for the Northern Hemisphere. Using an epidemiological model (SEIR) of season flu in the US, we evaluated the impact of updating vaccine decisions on the epidemic size and the number of flu hospitalizations in the United States.

**Results:** In the past flu seasons between 2012 and 2020, the timing when the clades or subclades that predominantly circulated during flu season emerged varied by season. In particular, in 2013/14, season-dominant H3N2 subclade emerged after vaccine decisions were made, contributing to the mismatch between vaccine and circulating virus. If the H3N2 component of the vaccine were updated given the additional viral activity data collected after February, our simulation model showed that the updated vaccine could have averted 5,000-65,000 flu hospitalizations, depending on how much vaccine effectiveness could improve with matching vaccine virus. On the other hand, updating the B/Victoria vaccine component did not yield substantial change in flu burden in the 2019/20 season.

**Conclusions:** With rapid vaccine production, revising the timeline for vaccine selection can result in substantial epidemiological benefits, particularly at times when additional data help improve the vaccine effectiveness through better match between vaccine and circulating viruses.

## Introduction

Seasonal influenza, or flu, is a respiratory infection caused by varying strains of circulating influenza viruses. Each season, a range of A and B strains of influenza viruses circulate, causing 3-5 million severe illnesses around the world (1). In the United States between 2010 and 2020, flu caused millions of illnesses, 140,000 – 710,000 hospitalizations, and 12,000 – 52,000 deaths every season (2). Influenza vaccine has played a key role in curtailing transmissions between individuals and reducing risks of severe complications from infections (3). According to the Center for Disease Prevention Center in the United States, flu vaccine reduces the risk of flu illness by 40-60% in the seasons when vaccine viruses well matched with circulating viruses (4). Vaccine effectiveness can vary season-to-season for multiple reasons. The characteristics of vaccinated people such as their age, prior exposure and vaccination history, and underlying health conditions can affect vaccine effectiveness (5, 6, 7). How well the viruses chosen for the seasonal flu vaccine resembles the viruses that will circulate during the flu season is another important determinant of vaccine effectiveness (8, 9, 10, 11, 12, 13) Because the influenza virus continuously evolves through mutation in viral proteins (i.e. antigenic drift) or through reassortment between two different viruses (i.e. antigenic shift), it is challenging to predict circulating virus ahead of flu seasons(14).

To consolidate a global effort to respond to emerging variants and formulate adequate seasonal flu vaccines, in February and August each year, the World Health Organization (WHO) and its collaborating centers analyze the clinical specimens collected through the global surveillance system and select the influenza strains that are most likely to dominate the coming influenza seasons in the Northern and Southern Hemispheres, respectively (15). The entity carefully monitors the trend in viral activity observed over the past season up to the decision-making points and considers the genetic and antigenic characteristics of new variants, prior to selecting the candidate vaccine for the following season. The current timeline for seasonal flu vaccine decision and formulation is set mainly to ensure that after vaccine candidate is shared with manufacturers, 450-500 million fertilized chicken eggs are ready to grow virus so bulk-production of the vaccine finishes in time for distribution, given that most licensed seasonal flu vaccines have been made in eggs (16). However, deciding vaccine candidates 5-6 months ahead of the flu season allows time for the circulating virus to evolve from the selected vaccine strain, lowering the chance of matching vaccine strain to circulating strain and potentially reducing vaccine effectiveness.

The next generation of flu vaccines whose manufacturing process does not rely on egg supply has emerged with the potential to shorten the vaccine production time and to improve vaccine efficacy. This advancement has made the potential for mass production of flu vaccines within 1–4 months, allowing time for collecting additional flu specimens from the WHO surveillance system to detect changes in viral activities and to identify emerging viruses before choosing vaccine candidate strains for the next flu season. Cell-based vaccine, for example, substitutes mammalian cells for chicken eggs to grow candidate vaccine viruses, which can be mass-produced and stored ahead of the season (17). A new recombinant technology that genetically engineers a vector virus so that it expresses the antigenic proteins in vector virus-infected insects or plants can produce large amounts of flu vaccines in 5–8 weeks (18). Since COVID-19 pandemic, mRNA vaccines start to expand to other respiratory diseases including seasonal flu, with their capability to deliver antigenic proteins without replicating virus in cells or eggs (19, 20).

The potential benefit of the next generation of seasonal flu vaccines have been reviewed in only a few studies in respect to improving vaccine effectiveness as a result of avoiding egg adaptation and expediting production process(17, 21, 22). However, there is little discussion as to whether the next generation of seasonal flu vaccines might revise the current vaccine decision and formulation schedule and its potential impact on the future seasonal flu epidemics. Systematic assessment of the past vaccine decisions given the observed trend in flu variants and evaluating the opportunity costs of early vaccine decision will provide insights to this question. Therefore, in this study, we aim to 1) identify the past flu seasons where the opportunity to improve vaccine decision existed if rapid vaccine production were available and 2) quantify the impact of revising the vaccine preparation and manufacturing schedule, where new vaccine production technologies allow more time for specimen collection prior to vaccine virus selection.

## Methods

### Seasons that additional data could have informed vaccine decision

We first investigated if the past decisions for seasonal flu vaccine between 2012 and 2020 could have changed if vaccine strain were chosen later than February for the Northern Hemisphere. Given that the level of viral activity and antigenic characteristics of emergent virus is key information in determining vaccine strain, we extracted data on the frequency of existing or emerging viruses over time from GSAID (23). In the GSAID’s genomic surveillance data, the frequency data were available every six months, in January and July. Given limited data frequency, we assumed that the genomic distribution of circulating virus in January represents the viral activity around the time when vaccine strain was chosen for the Northern Hemisphere. On the other hand, we considered the viral activity detected in July as potential data that could have been collected and used if the seasonal flu vaccine virus were chosen later than February. The viral activity of each clade and subclade in January of the following year indicated predominant and other co-circulating clades and subclades during the flu season. By examining the change in viral activity and the emergence of antigenically new virus between the three data points, we identified the seasons where the antigenically distinct viruses that predominantly circulated during the seasons were not selected due to their low activity in January but presented increasing activity before the season started in September of that year.

### Epidemiological impact of alternative flu vaccine decisions

In the flu seasons where the additional data collected after February potentially could have been used to update vaccine components, we quantified the epidemiological outcomes of the alternative vaccine decision in the US using an influenza transmission model. In our previous study, we built a mathematical model that simulates the seasonal flu epidemics from 2009 to 2020 in the US population (24). In brief, the non-age-stratified SEIR model simulates the co-circulation of four influenza subtypes including A(H1N1), A(H3N2), B(Yamagata), B(Victoria) co-circulate. The simulated population builds immunity against a specific flu strain based on their immunity history in the prior season and vaccination in the current season. The model considered vaccine uptake and strain-specific vaccine effectiveness that varies by season. Our previous study validated the model against the subtype distribution (i.e. percentage of flu cases attributed to each flu subtype) and seasonal flu hospitalization observed in the US from 2012/13 to 2019/20 season. Because the model was validated for the seasons later than 2011/12 and given the limited data on genomic surveillance data prior to 2012, our study focused on the seasons between 2012 and 2020.

In this simulation model, we compared the epidemiological outcomes in two scenarios: (1) historical flu seasons with the chosen vaccine strain and the observed vaccine effectiveness (2) flu seasons with alternative vaccine scheduling where extended time of data collection led to selecting candidate vaccine viruses that better match with circulating viruses and consequently improving vaccine effectiveness. An alternative vaccine decision considered updating one or multiple vaccine subtype compositions, depending on the viral activity and antigenic characteristics observed over the extended period.

In order to determine how much the vaccine effectiveness can improve in the later scenario, we first inferred the vaccine effectiveness against the antigenically similar (or ‘matching’) strains and effectiveness against antigenically dissimilar (or ‘mismatching’), assuming that the observed vaccine effectiveness is the weighted average of the two vaccine effectiveness (perfectly matching, mismatching). The prevalence of circulating viruses that match or mismatch with vaccine strain was the weight for the average.

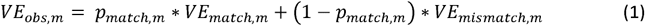

*(VE*_*obs*_,_*m*_ : observed vaccine effectiveness for subtype m,

*VE*_*match*_, _*m*_: vaccine effectiveness against the flu subtype m that are antigenically indistinguishable from the vaccine virus

*VE*_*mismatch*_, _*m*_, : vaccine effectiveness against the flu subtype m that are antigenically distinct from the vaccine virus

*P*_*match*_, _*m*_, : proportion of circulating viruses that are antigenically indistinguishable from vaccine viruses for subtype m)

Vaccine effectiveness with a perfectly matching vaccine strain is higher than when vaccine does not match with the circulating strain by a certain ratio (*rVE*).

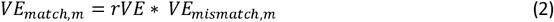

We then solved the equation (1) for *VE*_*mismatch*_,_*m*_, and hence *VE*_*match*_,_*m*_,. Estimated, and *VE*_*mismatch*_,_*m*_,, given *rVE* and *P*_*match*_, _*m*_, were presented in Table S1 and S2

In the scenario of historical flu season with original vaccine decision, we assumed that the prevalence of circulating viruses that match with the vaccine virus is equal to the percentage of circulating virus that were can be inhibited by antisera raised against the considered vaccine strain reported through the antigenic analysis performed by the Centers for Disease Control and Prevention (25). In contrast, in the scenario of alternative vaccine decision, we assumed that the circulating viruses that used to mismatch with the historical vaccine strain will match well with the updated vaccine strain. For example, in the historical scenario if the prevalence of matching and mismatching circulating viruses is 30% and 70%, the alternative vaccine decision would have 70% and 30% as the prevalence of matching and mismatching circulating viruses, respectively. We summarized the estimated subtype-specific vaccine effectiveness in those two scenarios in Table 1.

**Table 1.** Estimated vaccine effectiveness with original vs. updated vaccine strain selection in 2014/15 and 2019/20 season. rVE: the ratio in vaccine effectiveness against matching vs. mismatching circulating virus

| Season | 2014/15 season |  |  |  | 2019/20 season |  |  |  |
| --- | --- | --- | --- | --- | --- | --- | --- | --- |
| Vaccine decision | Original vax | Updated vax (rVE=1.28) | Updated vax (rVE=2) | Updated vax (rVE=10) | Original vax | Updated vax (rVE=1.28) | Updated vax (rVE=2) | Updated vax (rVE=10) |
| A(H1N1) | 0.30 | 0.30 | 0.30 | 0.30 | 0.30 | 0.30 | 0.30 | 0.30 |
| A(H3N2) | 0.06 | 0.07 | 0.09 | 0.19 | 0.30 | 0.30 | 0.30 | 0.30 |
| B(Yam) | 0.55 | 0.55 | 0.55 | 0.55 | 0.45 | 0.45 | 0.45 | 0.45 |
| B(Vic) | 0.55 | 0.55 | 0.55 | 0.55 | 0.45 | 0.50 | 0.59 | 0.89 |

We compared the number of infected individuals and the number of flu hospitalizations in the two scenarios to estimate the number of averted hospitalizations with the new vaccine scheduling in the past flu seasons where selected vaccines did not well match with circulating strains. We first estimated when the ratio in vaccine effectiveness for matching and mismatching circulating strain is 1.28, as estimated in a systematic review (26). Because how much vaccine effectiveness can improve when circulating strains match with vaccine strains, we performed sensitivity analysis by increasing the ratio from 1.28 to 2 and 10.

## Results

In the past flu seasons between 2012 and 2020, the timing when the clades or subclades that predominantly circulated during flu season emerged varied by season (Figure 1). The H1N1 clade that predominated the 2013/14 season, for example, showed increasing activity over the time from February in 2013 when vaccine decisions were made for the Northern Hemisphere to January in 2014 when the season has started. On the other hand, the B/Victoria clade that predominated when the vaccine decision was made for 2017/18 season showed decreasing activity over the same period.

**Fig 1.**
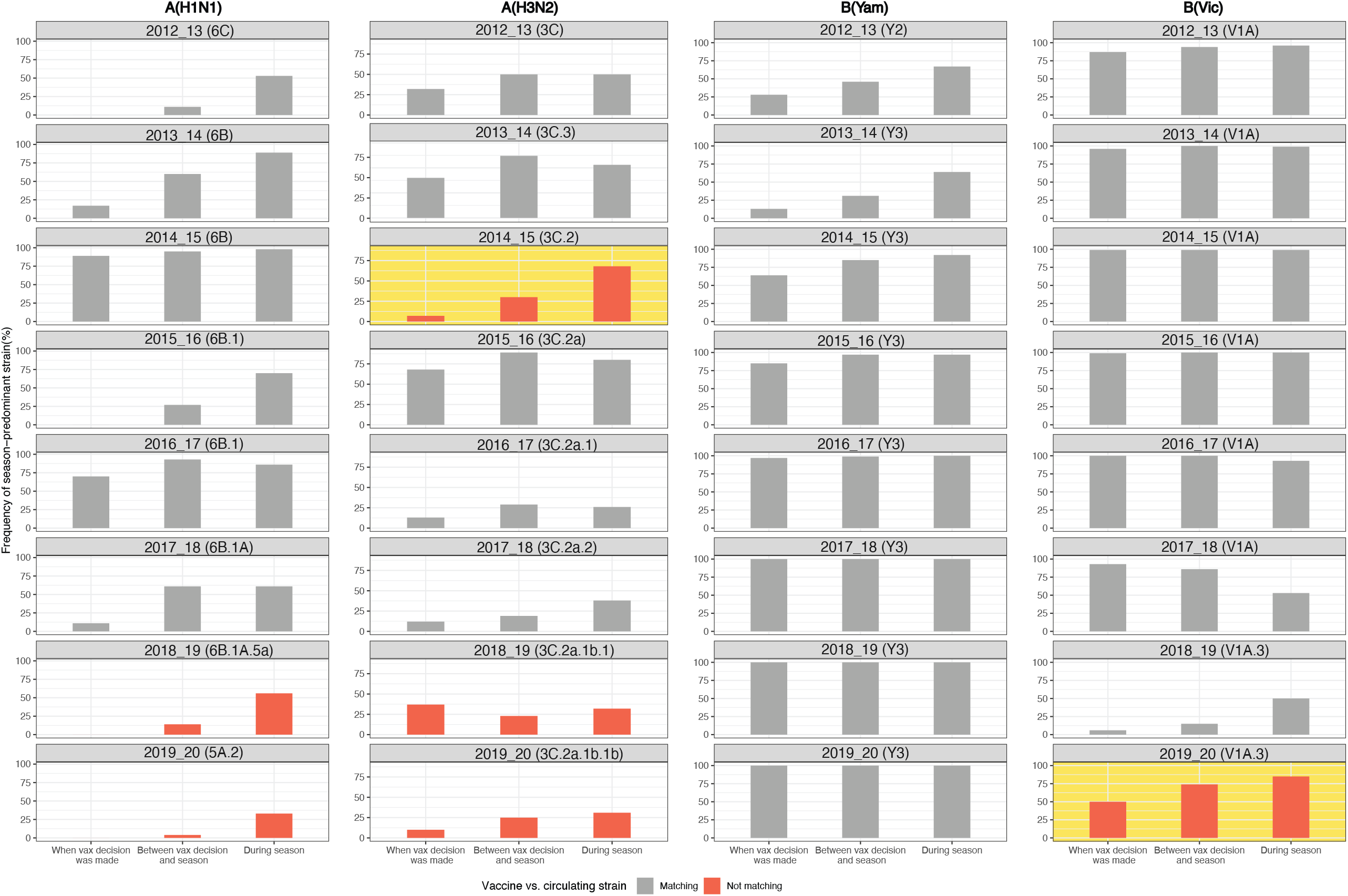
Trend in the frequency of season-predominant strain detected during the seasons between 2012/13 and 2019/20. The season-predominant strain of flu virus indicates the clade or subclade with the highest frequency detected during the season and was labeledIn each season, frequency was measured in three time points: when the vaccine decision is made (January prior season data), between vaccine decision and season (July data), and during season (January in season data). Bars show the trend in the frequency of predominant strains that are antigenically indistinguishable (gray, reduction in HI titre < 50%) and antigenically distinguishable (red, reduction in HI titre >= 50%) from vaccine viruses, respectively. Among the seasons with mismatching flu vaccines, influenza subtypes of which season-dominant strain presented higher than 25% frequency in July were highlighted in yellow as the vaccine components that could be potentially updated by delaying vaccine decision to July.

Among the clades or subclades that were predominant during the eight seasons, six were antigenically distinct from the selected vaccine virus. Four of these six viruses were present with very low activity (<10% frequency) when the vaccine decision was made. We found that the antigenically distinct and season-dominant H3N2 subclade in 2014/15 and B/Victoria clade in 2019/20, had presented greater than 25% activity during the time between vaccine decision and the flu season start, indicating that the vaccine decision could have changed for these subtypes if the decision was made later than February. In contrast, 6B.1A.5a subclade of H1N1 virus, which predominated during the 2018/19 season and was antigenically distinct from the H1N1 candidate virus, presented increasing activity only after the season started, exemplifying the case where delaying vaccine decision may have not yielded potential benefits.

In comparison to the past flu seasons with original vaccine decisions, we found that the alternate vaccine decision for A(H3N2) led to a decrease in the number of infections at the peak of the 2014/15 season (Figure 2A). If the ratio of vaccine effectiveness for matching virus compared to mismatching virus is assumed to be as high as 10 (rVE=10), we estimated even greater reduction in the number of infections at the season’s peak. Although the number of infections attributed to A(H3N2) decreased with the improved vaccine effectiveness for A(H3N2), the number of infections with other flu subtypes such as A(H1N1) increased (Figure 2B). In the scenario of updating the B(Victoria) vaccine component in the 2019/20 season, improved vaccine effectiveness for B(Victoria) did not result in significant change in the size of the flu epidemics in the 2019/20 season. In fact, when greater improvement in vaccine effectiveness with matching vaccine virus was assumed (rVE=10), updating the B(Victoria) vaccine component resulted in slightly more infections (Figure 2A). In this scenario, we found that the number of averted infections with improved B(Victoria) vaccine component was offset by the increase in the number of infections with other subtypes such as A(H1N1).

**Fig 2.**
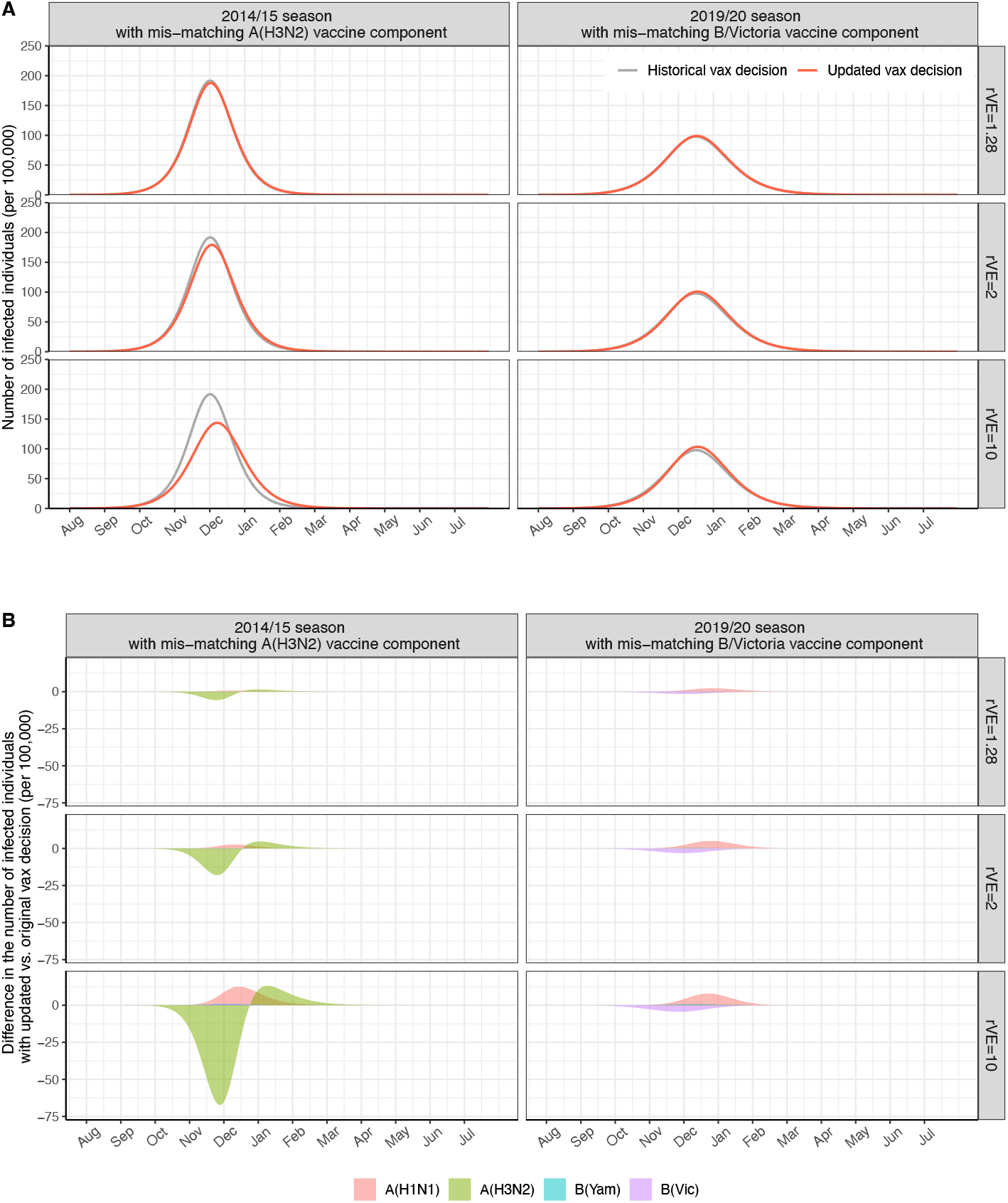
Historical flu epidemics vs. simulated flu epidemics with alternative vaccine decisions. Compared to the vaccine effectiveness in historical flu seasons, alternate vaccine decisions in 2014/15 and 2018/19 seasons led to higher vaccine effectiveness. In these scenarios, the ratio in vaccine effectiveness (rVE) between matching and not-matching vaccine virus varied as 1.28, 2, and 10.

**Fig 3.**
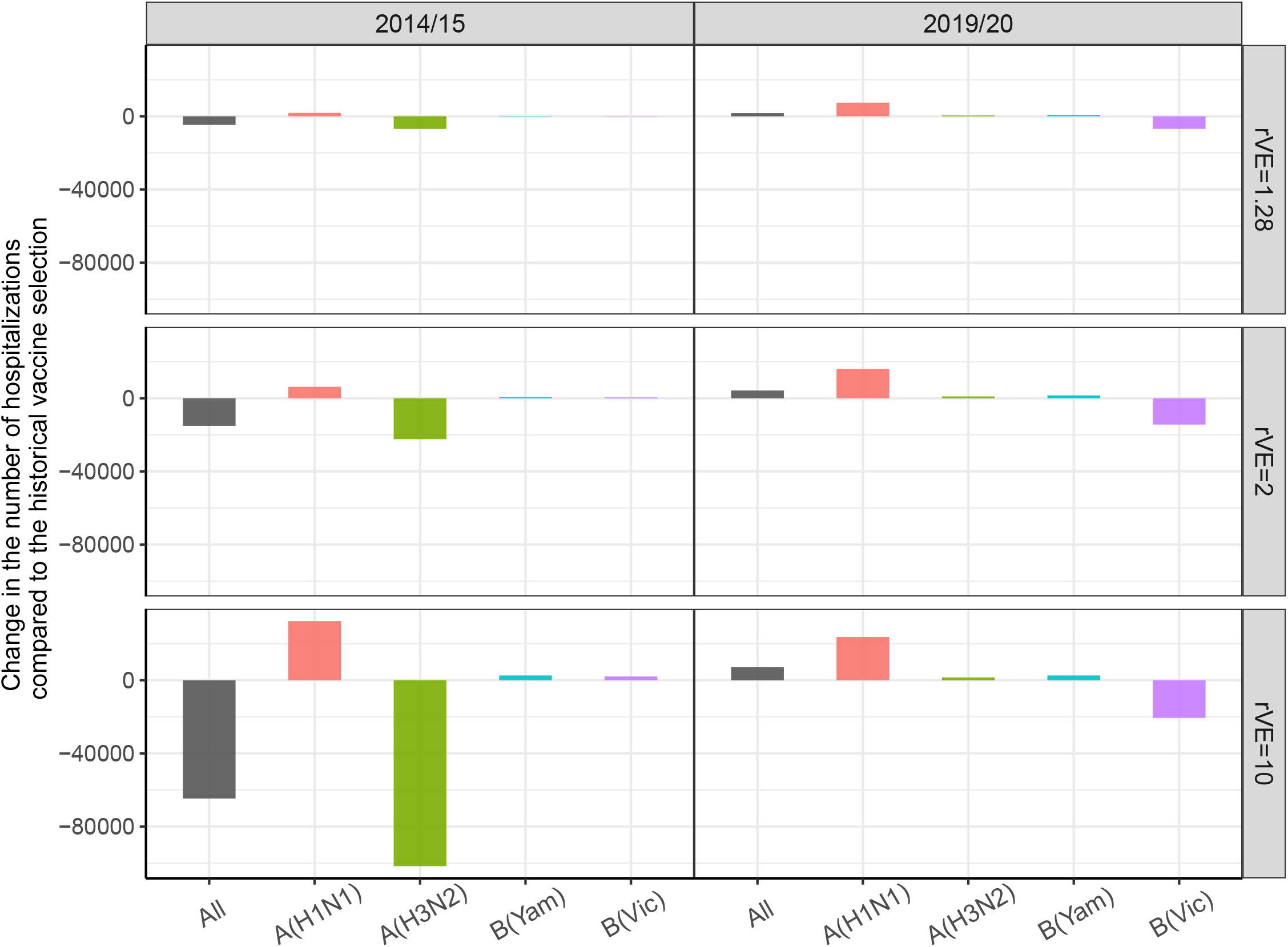
Number of flu hospitalizations that were averted with alternative vaccine decisions in 2014/15 and 2019/20 seasons. Ratio in vaccine effectiveness (rVE) between matching and not-matching vaccine virus varied as 1.28, 2, and 10.

Compared to the historical A(H3N2) vaccine component, the updated A(H3N2) vaccine resulted in 5,000 fewer flu hospitalizations in the 2014/15 season if the ratio in vaccine effectiveness between matching and mismatching vaccine virus is 1.28. If matching vaccine virus can achieve vaccine effectiveness 10 times effectiveness of mismatching vaccine (rVE=10), we estimated even greater number of averted hospitalizations by updating A(H3N2) vaccine virus, as high as 65,000 hospitalizations in the 2014/15 season. The subtype-specific analysis showed that this estimate is the combination of decrease in hospitalization attributed to A(H3N2) and increase in hospitalization due to other flu subtypes of which vaccine effectiveness was not updated. Similar to the previous result, updating B(Victoria) with the additional surveillance data resulted in increase in the total number of flu hospitalization. When we broke down the change in the number of hospitalizations by flu subtypes, we found that the increase in the number of hospitalizations in subtypes other than B(Victoria) was greater than the decrease in the number of hospitalizations due to B(Victoria) with the updated B(Victoria) vaccine composition.

## Discussion

Our study investigated the potential benefit of revising the current selection and formulation schedule for the next generation of influenza vaccine, by systematically evaluating the past influenza vaccine decisions based on the knowledge of viral circulation at the time of decision-making. Out of eight flu seasons from 2012 to 2020 in the Northern Hemisphere, we identified two seasons where antigenically new variants of flu virus emerged after the vaccine decision was made and their increasing activity was detected in the surveillance system before seasons started. In February of 2014, WHO recommended no change in the influenza A(H3N2) vaccine strain for 2014-15 season, based on the observation that H3N2 viruses detected in the surveillance system were antigenically similar to the 2013/14 vaccine reference virus (A/Texas/50/2012). However, the proportion of these drift variants increased starting from May, 2013, accounting for more than 80% of H3N2 virus detected in the Flu VE network(27, 28). Similarly, genetic testing of circulating virus prior to the 2019/20 season showed increasing diversity in the circulating B/Victoria virus (29). A vast majority of the emergent viruses had a three amino acid deletion in HA1 and were poorly prevented by the recommended B/Victoria component of flu vaccine. These two seasons exemplify the case where additional data collected after February could inform the selection for vaccine strain and improve the matching of the recommended vaccine strain to the circulating strain. On the other hand, we found that antigenic characteristics of new variants and the timing of when new variants emerged were highly variable in the past, emphasizing the challenges faced in improving vaccine effectiveness. For instance, an antigenic variant of H3N2 (3C.3a) emerged during the 2018/19 season and the antigenic difference between the circulating strain and the 2018/19 H3N2 reference virus (3C.2a1) was inevitable. Although detected increase in antigenic variants near season starts still does not guarantee its predominant activity during the season, the data collected over the extended period can be used in projecting the direction of viral evolution, which can also help predict the circulating virus (30).

Additional samples over the extended period before the season start can improve vaccine effectiveness and consequently reduce the flu burden. The epidemiological impact of alternative vaccine selection and production schedule depends on many factors including how much vaccine effectiveness can improve if vaccine strains match with circulating flu virus and how predominant the influenza subtype will be during the season. Our simulation showed that updating A(H3N2) vaccine component of 2014/15 season in the Northern Hemisphere resulted in substantial decrease in the number of hospitalizations in the US if the match between vaccine and circulating virus plays key role in improving seasonal flu vaccine effectiveness. However, updating B/Victoria in 2019/20 did not yield a similar impact even if high relative vaccine effectiveness is assumed. The difference in the estimated impact of updating vaccine component between the two seasons resulted in part from the fact that A(H3N2) accounted for 83% of flu cases in 2014/15 whereas B(Victoria) accounted for only 36.5% of flu cases in 2019/20. In contrast, improved vaccine component targeting the less-predominant subtype such as B(Victoria) in 2019/20 led to more infection by other flu subtypes that predominated the season (31, 32). This result highlights the importance of improving vaccine effectiveness, especially for the subtypes that are more likely to predominate in the season, such as A(H1N1) and A(H3N2) that predominated the historical flu seasons (25). The improvement in vaccine effectiveness with matching vaccine virus could be even higher in the next generation of flu vaccines by avoiding unnecessary change in vaccine strain in the progress of growing virus in eggs (33). Although the benefit of improved vaccine with matching vaccine virus is promising, other factors can affect the vaccine effectiveness including host factors (34)

In the past, WHO postponed selection for A(H3N2) component by one month in 2003 and 2019 in order to collect additional information on the emergent flu viruses (35). The next generation of flu vaccines, without the need for mass production of chicken eggs, allows even more time to perform data collection and antigenic characterization of emerging flu viruses before selecting vaccine candidate virus. For example, mRNA can complete mass production within a few weeks (20), potentially reducing even further time between vaccine decision and season start. On the other hand, the proposed timeline with delayed decision on vaccine strain entails the risk of delaying the supply and delivery of influenza vaccine. The flu vaccine supply shortage due to license suspension of a major vaccine manufacturer in 2004 showed that vaccine supply can be disrupted by many factors, which underscores need for careful examination of risks and benefits of revising vaccine formulation schedules (36). Even after vaccine strain is determined and vaccines are formulated, filled, and packed, the remaining steps for regulatory approval by FDA, vaccine release, and distribution can take 3-4 months (37). Potential delays in availability of flu vaccine may result in missed opportunities for vaccination resulting in reduced vaccine uptake. Hence, the benefits of delaying strain selection need to be balanced with providing adequate and timely flu vaccine supply.

This study quantifies the modeled impact of delaying flu vaccine selection but has limitations. First, the study inferred the trend in emerging flu virus based on the limited data available of two time points 6 months apart. We assumed that the decision for vaccine strain is made based on the increasing activity of emergent flu virus that was observed between two data points (January, July). Because it is less likely that vaccine decision can be postponed until July for the Northern Hemisphere, our study estimated the impact of updating vaccine decision in the best scenario. In addition, decision makers may have access to more granular information on viral trends. Delaying vaccine decisions to collect more data could lead to different decisions on vaccine strains. Availability of granular information on the perhaps monthly activity trend in combination with genetic and antigenic characteristics will offer the opportunity to explore optimal timing of making flu vaccine decisions in future studies. Second, in estimating the impact of alternative vaccine decisions with better match to circulating strains, we assumed that flu vaccine with antigenic similarity to circulating strain will present higher vaccine effectiveness than one with antigenic dissimilarity, without specifying vaccine types. However, the relative vaccine effectiveness with matching vaccines can vary by vaccine type as well as age and vaccination history of the infected population (34, 38). We performed sensitivity analysis on the relative vaccine effectiveness to address this limitation. Lastly, we assumed that antigenic similarity determines the vaccine effectiveness, but vaccine effectiveness may not consistently be associated with the degree of antigenic similarity as measured by the hemagglutinin inhibition assay (39).

As new and innovative types of flu vaccine emerge with the potential for improving speed and efficacy of production and vaccine effectiveness, the current formulation, production, and regulatory process for seasonal flu vaccine needs to be carefully reassessed. Our study investigated the potential benefit of having later vaccine strain decisions and collecting data over an extended time, which could be feasible with the next generation of flu vaccines. Our study concluded that revising the timeline for vaccine selection can result in substantial epidemiological benefits, particularly at times when additional data help improve the vaccine effectiveness through better match between vaccine and circulating viruses.

## Data Availability

All data produced are available online at https://gisaid.org

## Notes

## Acknowledgements

We thank Dr.Flannery (US Centers for Disease Control and Prevention) for comments on the manuscript. We also thank Dr.Zimmerman (University of Pittsburgh) for providing suggestions in the early stage of research development.

## Ethics approval

As all data used in this study were publicly available, ethics approval was not required.

## Patient Consent Statement

our study does not include factors necessitating patient consent.

## Data availability

all the data used for this analysis are publicly available. Data from the Center for Disease Control and Prevention for influenza are available at https://www.cdc.gov/flu/index.html.

## Appendix

**Table S1.**
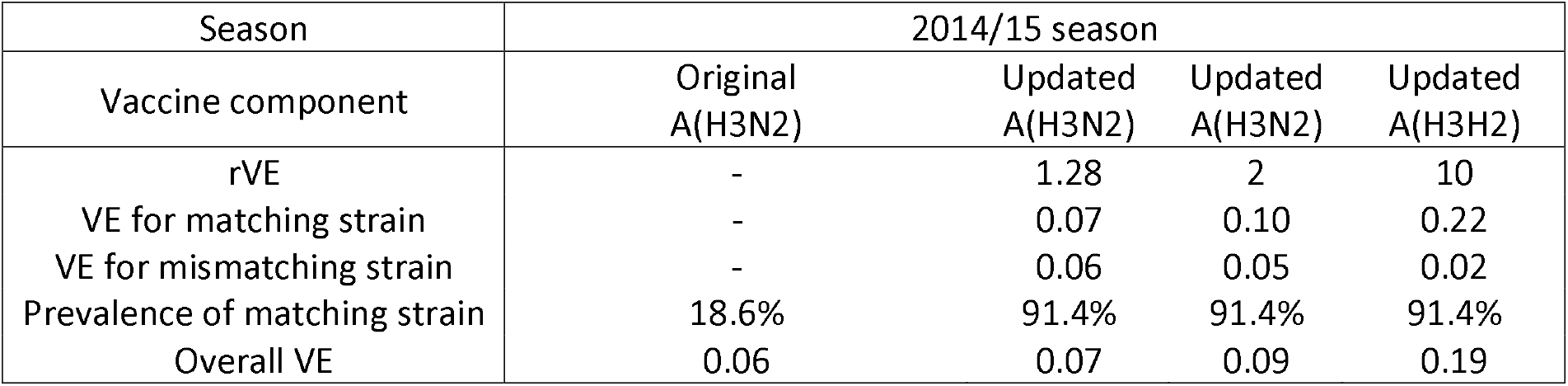
Vaccine effectiveness and prevalence of matching and mismatching strain under the scenario of original vs. updated vaccine components in 2014/15.

| Season | 2014/15 season |  |  |  |
| --- | --- | --- | --- | --- |
| Vaccine component | Original<br>A(H3N2) | Updated<br>A(H3N2) | Updated<br>A(H3N2) | Updated<br>A(H3H2) |
| rVE | - | 1.28 | 2 | 10 |
| VE for matching strain | - | 0.07 | 0.10 | 0.22 |
| VE for mismatching strain | - | 0.06 | 0.05 | 0.02 |
| Prevalence of matching strain | 18.6% | 91.4% | 91.4% | 91.4% |
| Overall VE | 0.06 | 0.07 | 0.09 | 0.19 |

**Table S2.**
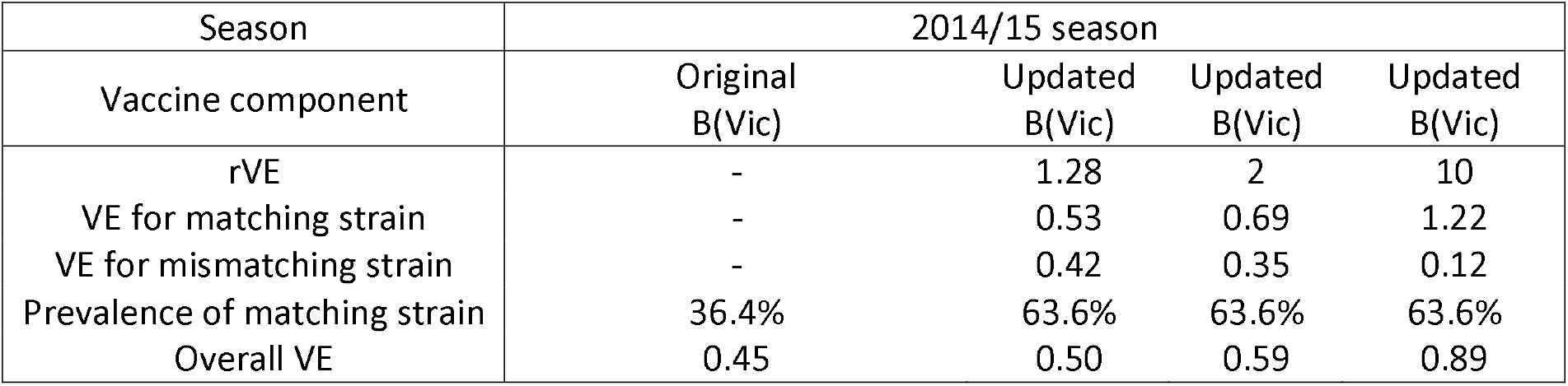
Vaccine effectiveness and prevalence of matching and mismatching strain under the scenario of original vs. updated vaccine components in 2019/20.

